# Last Mile elimination activities in Cambodia, October 2019 – December 2023

**DOI:** 10.64898/2026.05.12.26353080

**Authors:** Elijah Filip, Siv Sovannaroth, Alexa Kugler, Hannah Brindle, Pengby Ngor, Bunmeng Chhun, Pascal Ringwald, Zaixing Zhang, Rekol Huy

## Abstract

Between 2015 and 2025, Cambodia reported a 99.9% decline in the number of cases of malaria. To aid acceleration of elimination, the National Center for Parasitology, Entomology and Malaria Control (CNM) implemented a package of interventions known as the Last Mile (LM) elimination program. The aim of this study was to determine the impact of the LM program on case numbers and evaluate the coverage of interventions. LM was rolled out between November 2020 and December 2023 in villages reporting a locally acquired case of *Plasmodium falciparum* or mixed infection with *P. falciparum* and *P. vivax* and included combinations of targeted drug administration (TDA), intermittent preventative treatment for forest goers (IPTf), active fever screening (AFS), the recruitment of a village or mobile malaria worker (VMW/MMW) and the top-up of insecticide-treated bed nets (ITN) depending on the vulnerability and receptivity of the village.

A total of 103 full and 82 partial villages in seven provinces were included. Two rounds of TDA were administered, with a total of 10,678 individuals (67.6%) given during the first round and 9,678 (62.3%) during the second round. Coverage varied by province with none meeting the recommended threshold of 80%. IPTf was implemented each month among 35% (n=35) of full LM villages and 56% (n=42) of partial LM villages. A total of 11.7% (n=12) of full LM villages implemented AFS consistently on a weekly basis. Controlled interrupted time series showed no statistically significant difference in the number of malaria cases before and after the implementation of LM.

Although we were unable to prove a statistically significant impact of LM, likely due to the small number of cases prior to LM, it is important to add to the limited evidence-based for Accelerator Strategies in countries approaching the elimination of malaria. Furthermore, findings from the feasibility and impact of individual interventions were used to change policy at the national level.

## Introduction

Cambodia has made significant strides in malaria control and elimination in recent years from 74,099 cases reported in 2015 to 107 cases in 2025, a 99.9% decrease. During 2025, 51% of the cases were internationally imported and no indigenous cases of *Plasmodium falciparum* have been reported since January 2024. This progress was almost impeded by a steep increase in cases reported between 2017 and 2018, during which 46,590 and 63,186 cases were reported, respectively [1]. This prompted the National Center for Parasitology, Entomology and Malaria Control (CNM), with support from the World Health Organization (WHO), to implement a series of targeted intervention packages aimed at quickly reducing the burden of malaria, in particular *P. falciparum*. These included two Intensification Plans (IP) and the Last Mile (LM) program.

The IPs were implemented from October 2018 to December 2020 in 45 health center catchment areas in seven provinces with the highest reported number of cases of malaria. The IPs included targeted distribution of long-lasting insecticide treated bednets, additional support and supervision from CNM to the subnational level, creation of mobile malaria workers (MMW) deployed to high-risk areas, often outside of formal villages, revision of the national treatment guidelines, and improvements to supply chain management [2].

Following the implementation of the IPs, cases in the country decreased steadily alongside large increases in the number of parasitological tests performed. In particular, *Plasmodium falciparum* cases declined by 97% from 762 cases in October 2018 to only 23 cases in November 2020 [1]. As the number of malaria cases declined case burden became increasingly focalized in only a few provinces, namely Kampong Speu, Kratie, Mondulkiri, Ratanakiri, and Stung Treng [1]. Based on this progress, Cambodia set targets for *P. falciparum* elimination by 2023 and elimination of all malaria species by 2025 as part of the Malaria Elimination Action Framework 2021-2025 (MEAF2), endorsed by Cambodia’s Ministry of Health and the prime minister [3].

To accelerate *P. falciparum* elimination in remaining hotspots which were often associated with transmission among forest goers, CNM and WHO implemented the LM focus response package between November 2020 and December 2023. The aim of LM was to detect, investigate, and clear all foci with sustained, local transmission of *P. falciparum* or mixed *P. falciparum* and *P. vivax* cases. In 2024, CNM, in partnership with the Clinton Health Access Initiative (CHAI), conducted an evaluation of the LM program to assess whether the strategy contributed to a reduction in malaria cases and document the challenges, best practices and lessons learned. Results from evaluation of implementation indicators and impact analysis are presented here with the following objectives:

1. To estimate the difference in the number of *P. falciparum* and *P. falciparum/P. vivax* mixed cases before and after the implementation of LM
2. To estimate the difference in the number of *P. vivax* cases before and after the implementation of LM.
3. To determine the coverage of interventions in LM villages.

## Methods

### Sites

LM was piloted beginning in November 2020 in Phnom Srouch and Kampong Speu operational districts (OD) in Kampong Speu province. Between January 2021 and December 2023, LM was scaled up across seven provinces: Kampong Speu, Kratie, Mondulkiri, Preah Vihear, Pursat, Ratanakiri, and Stung Treng. Villages which received the full package of last mile interventions were referred to as ‘full’ last mile villages, whereas those which received only a subset of the intervention package were referred to as ‘partial’ last mile villages. Once LM was initiated in a village, implementation continued until the program ended in December 2023.

### Interventions

Last Mile consisted of a package of response interventions. When a locally acquired case (L1) of *P. falciparum* or a mixed infection of *P. falciparum* and *P. vivax* was identified in a village it triggered a focus investigation to determine the receptivity and vulnerability of the village. The results receptivity and vulnerability scores informed the LM intervention package for that village (Figure 2).

Interventions included a combination of four core interventions: recruitment of a village or mobile malaria worker (VMW/MMW), targeted drug administration (TDA), intermittent preventive treatment for forest-goers (IPTf), and top-up of insecticide treated bednets (ITN). VMW or MMW were recruited to provide passive case detection if the focus did not already have one. ITN distribution included the top-up of long-lasting insecticide treated nets (LLINs) and continual distribution of long-lasting insecticide-treated hammock nets (LLIHNs) to high-risk populations. TDA and IPTf were both targeted to males aged 15-49 years, the demographic generally considered at highest risk of malaria infection in Cambodia. TDA was provided to all eligible individuals in the village, whereas IPTf was provided only to those who intended to work in nearby forested areas in the following month. TDA was provided to the target population on specific dates. Individuals were directly observed taking the first dose of the TDA drug and provided with two further doses to take on their own. For IPTf, only the first dose of treatment was directly observed with the second and third doses self-administered by forest-goers; in some instances, all three doses were self-administered. Both TDA and IPTf were initially implemented with artesunate-mefloquine (ASMQ). However, in 2022, the drug regimen was changed to artesunate-pyronaridine (Pyramax). In villages implementing IPTf, weekly active fever screening (AFS) was additionally conducted to all high-risk populations.

As the program progressed and the incidence of *P. falciparum*/mixed cases was low, the program was expanded to apply a subset of interventions to villages which reported ‘local Cambodia’ (LC) cases (equivalent to domestically imported cases). These villages were classified as ‘partial’ last mile villages but only received IPTf. The national malaria surveillance guidelines were revised in 2021 and implemented in 2022 to incorporate last mile activities into routine surveillance practices [4].

### Data

Data were initially recorded on paper forms and reported to the national program using Excel-based monthly reports. Modules for the LM program were created in Cambodia’s Malaria Information System (MIS) used for case notification. Data were retroactively entered into the MIS by subnational and CNM staff. This analysis utilized data from the MIS, accessed on 22 May 2024.

### Analysis

Data were extracted from CNM’s Malaria Information System (MIS) database and included routine surveillance case data and intervention data collected as part of LM. Analyses were performed using R version 4.3.0.

Service outcome and output indicators were evaluated using descriptive statistics. Timeliness of LM initiation was defined as the time between the last reported *P. falciparum* or mix case and the date LM activities were reported to have started considering the staggered start dates of different provinces. Completion, timeliness, and coverage of targeted drug administration was calculated for both rounds of TDA completion was defined as implementation of two rounds of TDA, timeliness as the time between rounds and coverage as the proportion of the target population which received the first dose of TDA in each round. The target population was determined during the village-wide household census conducted at the start of last mile focus investigation. Coverage targets of 80% were set based on preexisting literature on MDA and TDA strategies [5]. Similarly, monthly completion and coverage of IPTf was calculated for each village and only included the first dose of IPTf, which was administered through DOT. Weekly completion of AFS was calculated for each village, and total tests are reported. Expected number of AFS rounds is calculated as the number of weeks from the first round of AFS in the village until the last week of December 2023.

Controlled interrupted time series (CITS) analysis using a negative binominal model of monthly case data relative to the start date of LM activities was utilized to determine the impact of the LM response package on the level and trend in cases. A negative binomial model was chosen to account for overdispersion within the data. Additionally, the Akaike Information Criterion (AIC) was lower using negative binomial compared to Poisson models. Villages which began implementing LM after December 2022 were excluded to ensure comparison with data for at least 12 months post-intervention was possible. Among the full LM villages, only those which completed two rounds of TDA and had at least one case of the relevant species (*P. falciparum* or *P. falciparum*/*P. vivax* mixed, or P. vivax only) were included. The primary analysis focused on impact in relation to the number of *P. falciparum* or mixed cases, and secondary analyses considered impact on *P. vivax*. CITS compared the trend in malaria cases before and after LM implementation in full, partial, and non-LM (comparison) villages within the same commune. Cases were aggregated by village relative to the start of LM activities, including cases within 12 months before and after implementation. For non-LM villages, cases were aggregated relative to the start of LM activities within the commune.

## Results

### Implementation

Between January 2021 and December 2023, a total of 103 full and 82 partial villages were included in the LM program. The total number of villages per province ranged from seven (two full and five partial) in Pursat to 57 (39 full and 18 partial) in Mondulkiri (Figure 1). A total of 54,172 residents lived in 11,262 households in full LM villages and 25,156 residents in 6,996 households in partial LM villages.

**Figure 1.**
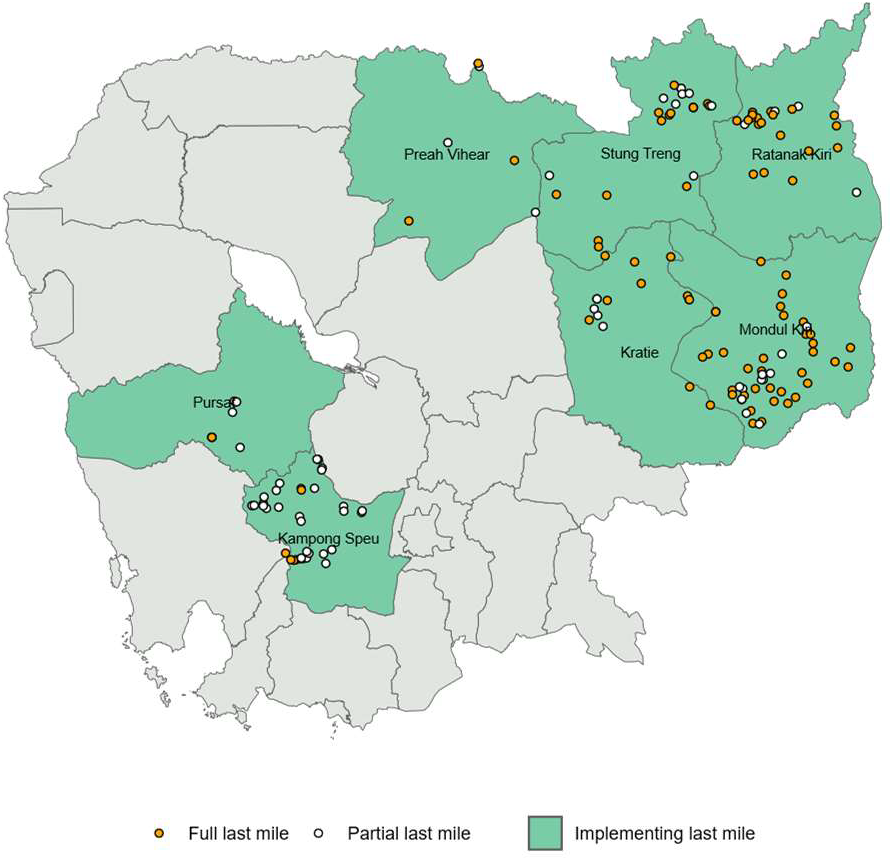
Location of full and partial Last Mile villages*. * 25 villages were excluded from the figure which did not have GPS coordinates.

**Figure 2.**
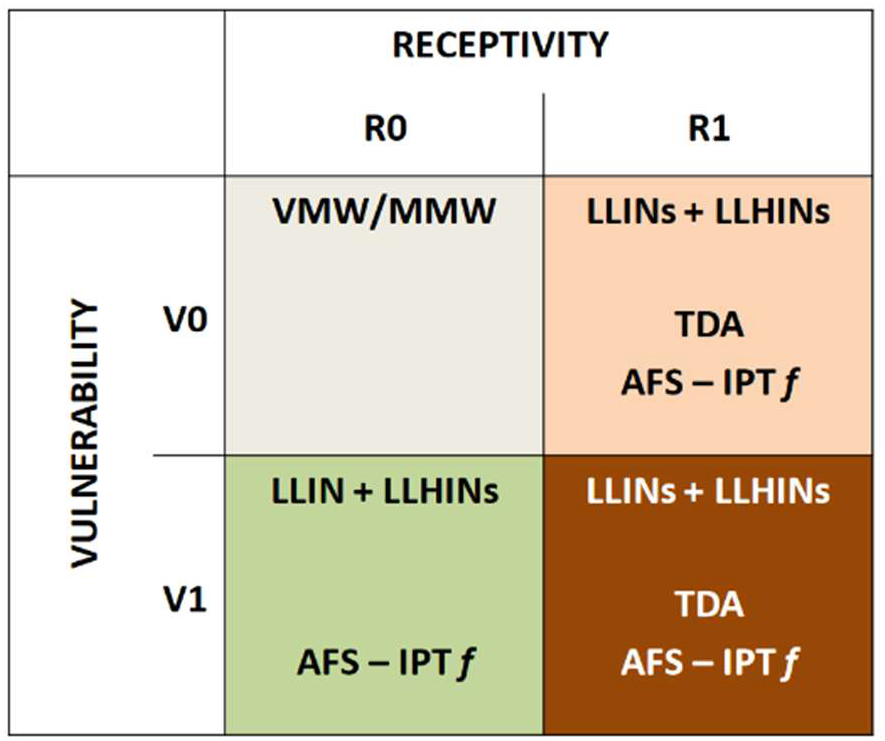
Last mile interventions according to village receptivity and vulnerability score.

Of the full LME villages which started LM activities following the most recent L1 *P. falciparum*/mix case (n=83), the median time for villages to start LM activities was 95 days [IQR=47.5-186 days]. This delay decreased with time. During 2021 the median time was 123 days [IQR=112-134 days] compared to 95 days [IQR=57-125 days] in 2023. A total of six villages implemented LM activities within 31 days.

### Interventions

#### Targeted Drug Administration

Of the 103 full LM villages which received two rounds of TDA, the median time between starting LM and the first round (n=87 villages who started TDA after eligibility) was 20 days [IQR 7-63 days] and 55 villages started TDA within 31 days. A total of 16,102 individuals were targeted for TDA of whom, 10,678 (67.5%) completed the first round and 9,678 (62.3%), the second. At the provincial level, the median percentage per village of those targeted who completed the first round ranged from 63.3% in Kampong Speu to 90.0% in Pursat; and the second round, 43.4% in Preah Vihear to 80.4% in Mondulkiri. Both Mondulkiri and Pursat reached the target threshold of median 80% coverage [5] for both rounds (Figure 4).

**Figure 4.**
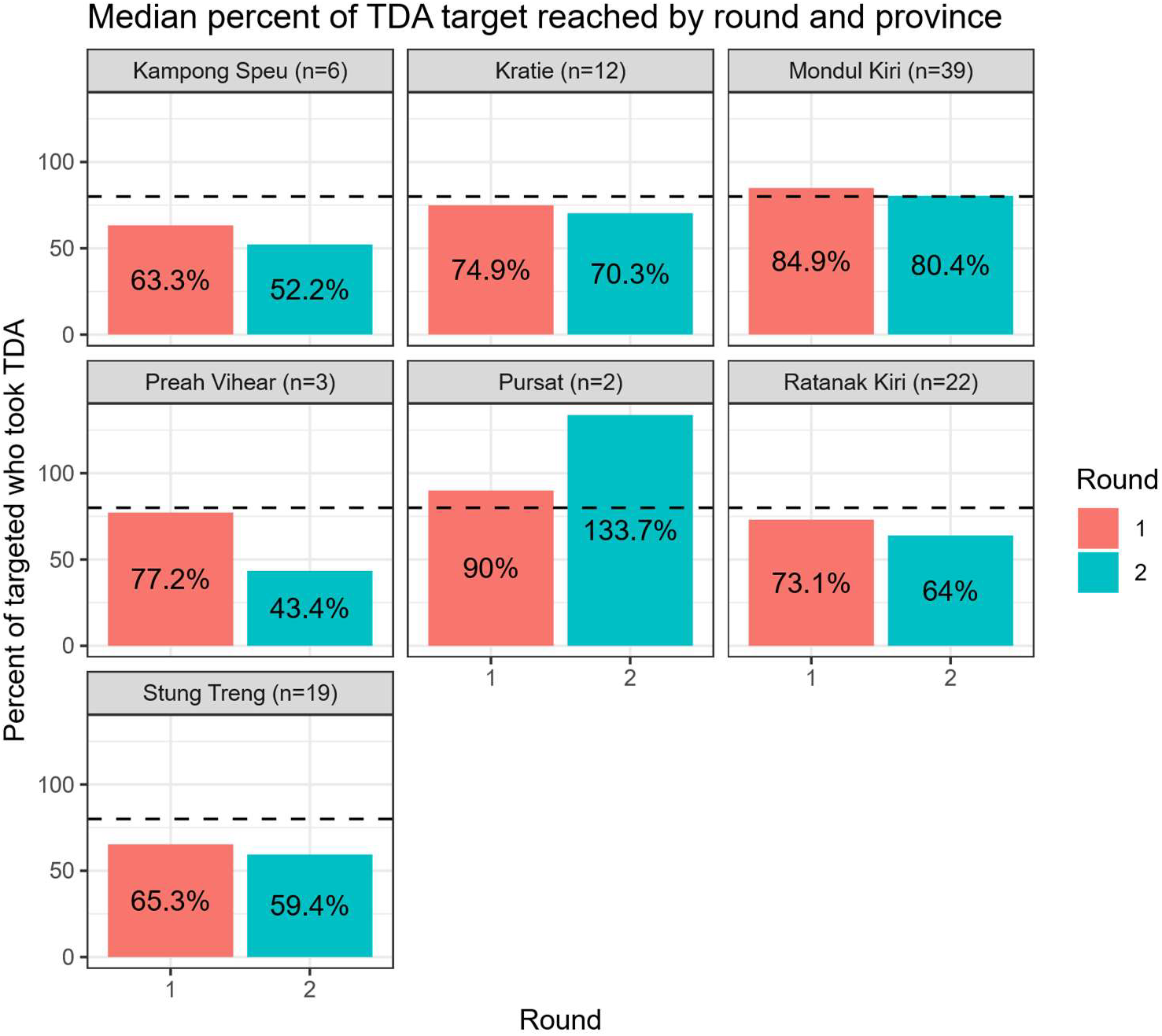
Median coverage of TDA round 1 and TDA round 2 by province. The number in brackets indicates the number of villages which implemented Last Mile activities.

#### Intermittent preventative treatment of forest goers (IPTf)

Of the 103 full LM villages, 100 implemented IPTf and 35% (n=35) implemented it each month since starting the activity. For the 82 partial LM villages, 75 implemented IPTf and 56% (n=42) implemented it each month.

The median coverage among forest-goers per village and month for the first month of IPTf was 14.3% [IQR 3.49-30.2%] and the second month, 20.0% [IQR 8.11-34.0%]. Among those villages implementing full LM activities, the highest median monthly coverage at the village level was reported in Kampong Speu (35.9%) and the lowest in Ratanakiri (8.3%). Among those villages implementing partial LM activities, the highest median monthly coverage was reported in Mondulkiri (30%) and the lowest, Stung Treng (16.2%) (Figure 5).

**Figure 5.**
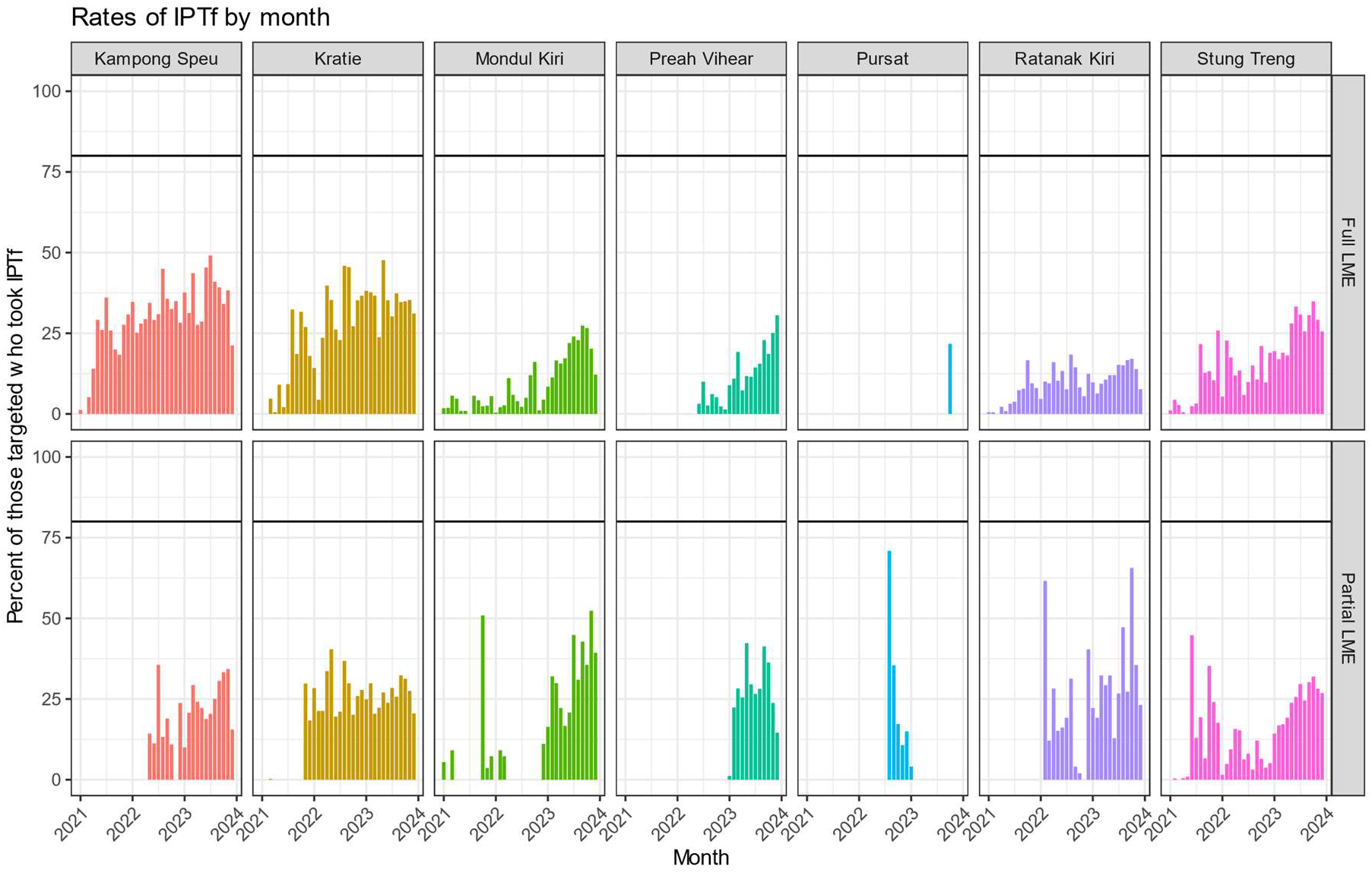
Monthly coverage of IPTf in full and partial Last Mile villages, by province.

#### Active fever screening

Active fever screening (AFS) was conducted in all 103 full LM foci, resulting in 221,121 tests. A median of 18 tests per village per week were performed [IQR 7-35]. This amounted to approximately 4% of the village population being tested per week (range, 2-8%).

A total of 11.7% (n=12) of full last mile villages implemented AFS consistently on a weekly basis (Figure 6). 58.3% (60/103) of villages implemented at least 80% of the expected number of AFS rounds. Of the provinces with foci that implemented at least 80% of the expected number of AFS rounds, Kampong Speu performed highest at 100% (6/6) of foci reaching this threshold, followed by Kratie at 75.0% (9/12). The highest median percentage of the residents per village tested per week was in Stung Treng (6.2%, IQR 3.2-10.5%) and lowest, Pursat (1.2%, IQR 0.3-6.4%).

**Figure 6.**
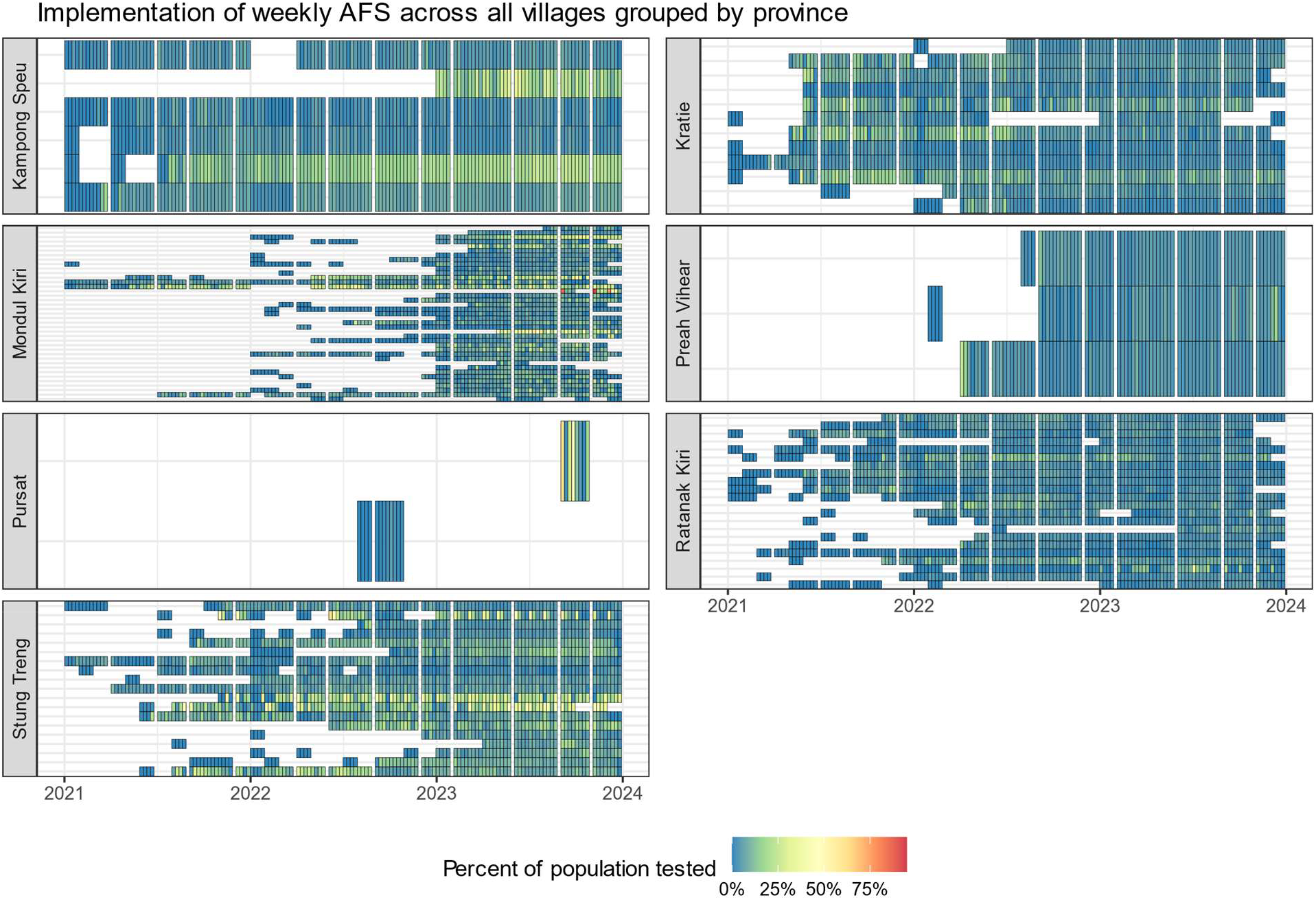
Implementation consistency of weekly active fever screening in full Last Mile villages.

### Controlled interrupted time series

For non-LM villages, those outside of communes with either full or partial LM villages were excluded.

#### Plasmodium falciparum and mixed cases

A total of 80 full LM villages and 381 comparison villages were included in the CITS to assess the impact of the full LM on *P. falciparum* and mixed cases (Figure 7.A). There were 22 *P. falciparum*/mix cases at the start of activities in full LM villages, decreasing to four cases by 12 months following initiation of LM. In non-LM villages, there were 14 *P. falciparum*/mix cases at the start of activities, decreasing to eight cases by 12 months.

**Figure 7.**
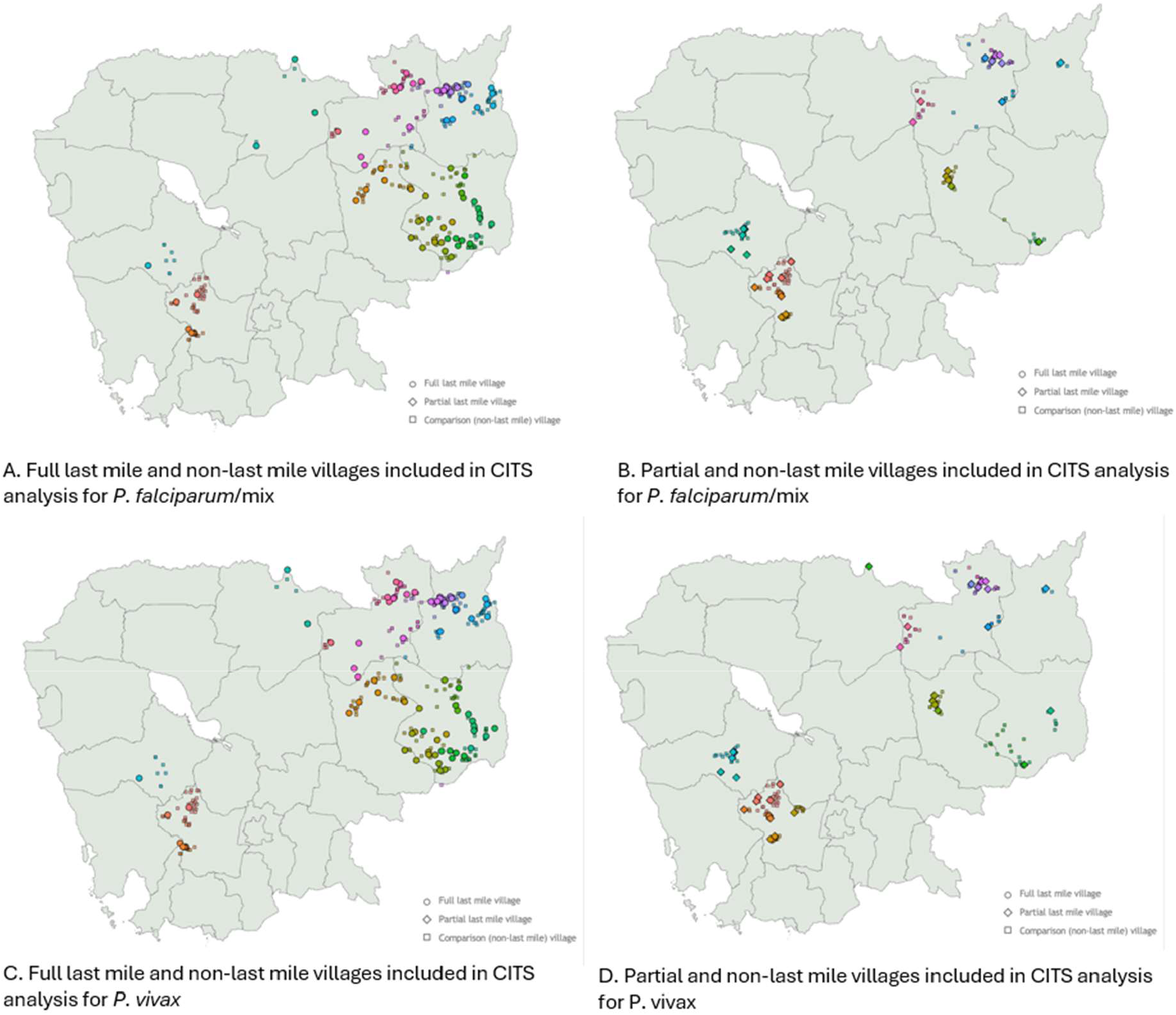
Location of full or partial Last Mile villages and comparison villages used for CITS analysis.

Controlling for these cases in non-LM villages, there was no statistically significant difference in the change in the number of cases (relative risk (RR)=1.425, p=0.577, 95% confidence interval (CI): 0.410 – 4.951) nor change in slope (RR=0.916, p=0.337, 95% CI=0.765 – 1.096) before and after the implementation of LM (Figure 8.A).

**Figure 8.**
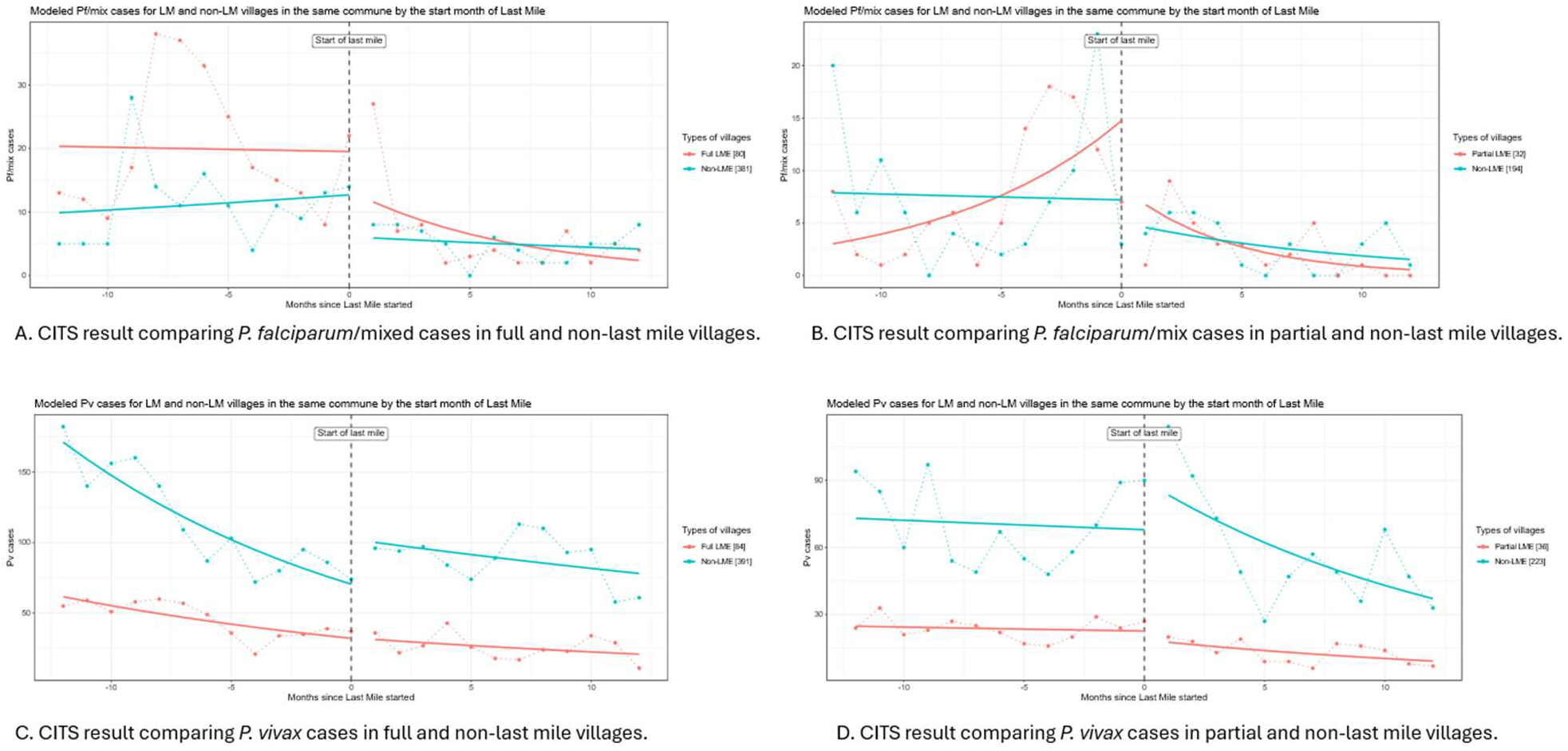
Controlled interrupted time series result of full last mile compared to non-Last Mile villages for *P. falciparum* (8.A) and *P. vivax* (8.C), as well as partial Last Mile compared to non-Last Mile (*P. falciparum*, 8.B; *P. vivax*, 8.D).

A total of 32 partial LM villages and 194 non-LM comparison villages were included in the CITS to assess the impact of the partial LM on *P. falciparum* and mixed cases (Figure 7.B). Among the partial LM villages, there were seven cases at the start of activities, decreasing to zero cases by 12 months. This is compared to three cases at the start of activities non-LM villages declining to one case by 12 months.

Controlling for *P. falciparum* and mixed cases in non-LM villages, there was no statistically significant difference in the change in the number of cases (RR: 0.819, p: 0.826, 95% CI: 0.137 – 4.882) nor in slope (RR: 0.765, p: 0.054, 95% CI: 0.583 – 1.005) before and after partial LM implementation (Figure 8.B).

#### Plasmodium vivax

A total of 84 full LM villages were compared to 391 non-LM villages within the same commune to assess the impact of LM on *P. vivax* cases (Figure 7.C). There were 37 *P. vivax* cases at the start of activities in full LM villages, decreasing to 11 cases at 12 months, compared to 74 *P. vivax* cases in non-LM villages, decreasing to 61 cases at 12 months.

Controlling for *P. vivax* cases in non-LM villages, there was no statistically significant difference in the change in the number of cases (RR: 0.695, p: 0.099, 95% CI: 0.452 – 1.070) nor the change in slope (RR: 0.967, p: 0.265, 95% CI: 0.911 – 1.026) (Figure 8.C).

A total of 36 partial LM villages were compared to 223 non-LM villages in the same communes to assess the impact of partial LM on *P. vivax* cases (Figure 7.D). There were 27 *P. vivax* cases at the start of activities in partial LM villages, declining to seven cases by 12 months, compared to 90 *P. vivax* cases at the start of activities in non-LM villages, declining to 33 cases by 12 months.

Controlling for *P. vivax* cases in non-LM villages, there was no statistically significant difference in the change in the number of cases (RR: 0.624, p: 0.144, 95% CI: 0.332 – 1.174) nor the change in slope (RR: 1.016, p: 0.722, 95% CI: 0.929 – 1.112) (Figure 8.D).

## Discussion

The World Health Organization (WHO) proposes recommendations for countries approaching the last stages of elimination including mass, targeted and reactive strategies which together may be classified as ‘Accelerator Strategies.’ [6] However, there are few publications which clearly outline their implementation and impact. Therefore, here we provide the methodology and analysis of the Last Mile program, an Accelerator Strategy approach used in Cambodia between 2021 and 2023, a year before no further indigenous *P. falciparum* cases were reported. This program of activities followed the Intensification Plan, a separate set of interventions implemented between 2018 and 2020 [7].

CITS analyses comparing villages which implemented LM activities compared to those which did not, did not find any statistically significant difference in the decline in case numbers of *P. falciparum* or *P. vivax*. While this might suggest that LM did not have any impact on case numbers compared to routine interventions such as passive case detection and treatment, vector control measures such as mass ITN campaigns and the utilization of pre-existing VMWs, the very small number of cases at the start of the implementation of LM and therefore insufficient power to detect any difference in the decline of these makes it challenging to infer any conclusions about the impact of the LM package. A power calculation was not performed prior to LM as this strategy was implemented for operational rather than research purposes, with the CITS analyses performed retrospectively. Additionally, the villages selected as controls may not have been at the same risk of malaria transmission as LM villages given that they did not report cases. Therefore, there were limitations in terms of comparability.

Despite this, the evaluation of individual interventions uncovered many interesting and useful findings. WHO provides a conditional recommendation for the use of TDA in areas with very low to low transmission or post-elimination settings with a very low certainty of evidence of the benefits and harms [8]. Our study was not designed to determine the impact of TDA as an individual intervention. However, WHO note that a critical aspect to the success of mass drug administration strategies is the achievement of high coverage (80% or greater) [5,8]. In Cambodia the coverage of TDA varied geographically and when calculating the median coverage per village, the threshold of 80% was only met in Mondulkiri for one of the two rounds. Previous accelerator strategies of the National Program (IP) did not include TDA or any form of mass drug administration, however, in a previous study of the impact of TDA using Pyramax among forest goers in Pursat, Cambodia during 2020 and 2021, the combined coverage of TDA and IPTf in targeted villages during round one was reported as 92% and during round two, 65% [9]. Coverage in the first round of this study was therefore higher than that reported in all provinces included in LM activities. Coverage during the second round was more comparable, except for Kampong Speu and Preah Vihear which reported lower coverage during LM. Given the small number of villages included in the analysis in these provinces, the results should be interpreted with caution. The authors of the study in Pursat noted that approximately one third of forest goers did not receive the second round of TDA as they claimed they were no longer visiting the forest [9]. While the results of key informant interviews which took place as part of LM activities (not included in this report) indicated that some forest goers had concerns about the side effects of AS-MQ or were reluctant to avoid drinking alcohol when taking either AS-MQ or Pyramax [10], it is unclear why we saw a large difference in coverage between rounds one and two. However, we recommend continuing to ensure that village leaders and other respected members of the community are involved in community engagement and TDA implementation. In addition, the effectiveness of TDA should continue to be evaluated where feasible, particularly as an accelerator strategy for *Plasmodium vivax* elimination, which is more challenging due to the presence of hypnozoites.

While there are no WHO recommendations for the use of IPTf, it has been used in recent years in Cambodia and Lao People’s Democratic Republic [9,11–14]. The coverage of IPTf as part of LM was far below the recommended threshold of 80% in all provinces and also that of the combined coverage of TDA and IPTf in the study in Pursat [9]. However, our results may have been subject to challenges with data quality. Unfortunately, the data reported to the MIS does not capture a dynamic denominator for the monthly number of forest-goers. Despite this, as IPTf was the only intervention used in partial LM villages we can possibly attribute the decline in cases, although the CITS was not statistically significant, it may be a useful intervention for future focus responses. Indeed, following a cluster of seven cases detected in Kampong Speu province during 2025, IPTf with chloroquine was implemented during a 60-day campaign which interrupted transmission with no subsequent resurgence of cases yet. For both TDA and IPTf, we were only able to calculate coverage based on the administration of the first dose of drug as subsequent doses were self-administered. This also meant that we were unable to determine adherence to the full course. However, since LM attempts to determine adherence have been made. Following the 60-day campaign in Kampong Speu, a rapid survey was conducted to assess self-reported adherence to the first round of IPTf in ten target villages with 94% of participants stating that they took all doses (unpublished data, CNM).

Similarly, while WHO provides a conditional recommendation for reactive case detection and treatment to reduce the transmission of malaria in elimination settings with a very low certainty of evidence, there are no recommendations on the use of AFS. AFS was also included in the study in Pursat as a precursor to the implementation of TDA and IPTf [9] and as part of the Accelerator Strategies in Lao PDR [15]. However, during LM, less than two-thirds of villages were able to reach the 80% threshold of implementation of AFS and we were unable to ascertain the number of positive cases detected during AFS as these were not linked to recorded cases in the MIS. Therefore, in future we recommend that a system is in place to identify cases detected during reactive or proactive case detection to understand the value of AFS. However, given that AFS is resource intensive and there was limited evidence of an impact on malaria transmission, it was recommended that it be removed as a focus response activity in Cambodia.

## Conclusion

The analysis of the LM activities in Cambodia has importantly contributed to the evidence-base for Accelerator Strategies. However, in elimination settings where case numbers are low it can be challenging to determine a statistically significant impact, as was demonstrated in our study. Despite this, we learned important lessons about the feasibility of the implementation of individual interventions which were used to inform changes to policy. Should case numbers allow, we would recommend further analysis to determine the potential individual impact of TDA, AFS and IPTf on the incidence of malaria and to disaggregate the CITS by demographic groups. We would also suggest an analysis of the impact of LM on the performance of passive activities at health centers and by VMWs which continued throughout.

## Data Availability

National malaria surveillance data are available at https://mis.cnm.gov.kh/. Additional data are available are upon reasonable request to the Director Office of the National Center for Parasitology, Entomology and Malaria Control (CNM), Phnom Penh, Cambodia at.

## Acknowledgments

We thank all the community members who were involved in Last Mile activities and colleagues at CNM and partners for their support in the implementation and provision of data for analysis.

## Ethics

As this work was considered to be operational, ethical approval was not deemed to be required and hence, not sought.

## Disclaimer

PR and ZZ are staff members of WHO. The authors alone are responsible for the views expressed in this publication, which do not necessarily represent the decisions, policies, or views of WHO.

